# Machine learning⍰based prediction of cardiovascular disease risk in Africa using WHO Stepwise Surveys: 2014-2019

**DOI:** 10.64898/2026.02.23.26346870

**Authors:** Wingston Felix Ng’ambi, Fatma Aziza Merzouki, Janne Estill, Olivia Keiser, Erol Orel

## Abstract

**Introduction:** Cardiovascular diseases (CVDs) are the leading cause of death globally, with rising burdens in Africa due to ageing populations, lifestyle changes, and poor risk factor control. Conventional risk scores developed in high-income settings often perform poorly in African populations. Machine-learning (ML) approaches offer potential to improve prediction by capturing complex, non-linear interactions among demographic, behavioural, and biological factors. This study applies ML models to WHO STEPS survey data to generate context-specific CVD risk predictions across 12 African countries.

**Methods:** We analysed data from 60,294 adults collected in WHO STEPS surveys between 2014 and 2019 across 12 African countries. Three ML models; Elastic Net logistic regression (LASSO), Random Forest (RF), and XGBoost (XGB); were trained to predict self-reported CVD outcomes. Data were split into training (80%) and testing (20%) sets with five-fold cross-validation. Feature selection used the Boruta algorithm, and model performance was assessed via accuracy, sensitivity, specificity, AUC, F1 score, and Brier score.

**Results:** Overall CVD prevalence was 5%. Hypertension emerged as the strongest predictor across all models, followed by alcohol-related harm. Tree-based models outperformed regression approaches and conventional clinical scores, with XGBoost achieving the highest discrimination (AUC=0.769), balanced accuracy (0.699), and calibration (Brier score=0.195). Predicted risk trajectories were smoother and more clinically plausible than Framingham or WHO/ISH scores, particularly across age, sex, and hypertension status. LASSO and Random Forest performed moderately, while conventional risk scores showed poor discrimination and marked miscalibration.

**Conclusion:** Machine-learning approaches provide accurate, context-specific cardiovascular risk prediction in African populations. By highlighting modifiable risk factors such as hypertension and alcohol-related harm, these models support targeted interventions aligned with WHO PEN, HEARTS, and SBIRT strategies. The African CVD Risk Prediction Tool translates complex data into actionable insights, offering a scalable platform for prevention-focused, equitable cardiovascular care across diverse African settings.

## INTRODUCTION

Globally, cardiovascular diseases (CVDs) account for 17.9 million deaths annually, making them the leading cause of death, and the risk of CVDs rises with age ^1 2 3 4^. The risk of CVDs increases with age and is linked to sedentary lifestyles, poor diets, smoking, and physical inactivity, contributing to rising rates of diabetes, hypertension, and obesity ^3 5^. The World Health Organisation (WHO) aims to reduce premature CVD deaths by 33% by 2030 ^6^. In Africa, an epidemiological shift suggests non-communicable diseases (NCDs) may surpass infectious diseases within a decade. Heart attacks and strokes cause most CVD-related deaths, with one-third occurring in individuals under 70 ^2^. Managing risk factors is crucial for improving health and reducing healthcare costs.

Machine learning (ML) models are increasingly gaining prominence in predicting health outcomes such as hypertension^7^, heart disease^8^, and diabetes onset and complications ^9^. Although classical methods like logistic regression and Fisher’s discriminant analysis have been utilised for health outcome prediction for a long time ^10^, ML models now offer improved computational power by analyzing complex datasets. Validated models like pooled cohort equations (PCEs) ^11^, Globorisk ^12^, World Health Organization/International Society of Hypertension (WHO/ISH) ^13^, Framingham risk score (FRS) ^14^, systematic coronary risk evaluation (SCORE) ^15^, and QRISK3 ^16^have long been used for CVD prediction. The commonly used ML models for predicting health outcomes include: simple models (i.e. logistic regression (LR), naive Bayes (NB) and K-nearest neighbor (KNN)); intermediate models (i.e. decision trees (DT), generalized linear mixed-model tree (GLMM tree)); ensemble models (i.e. extreme gradient boosting (XGBoost), random forests (RF), AdaBoost Classifier (AB), Extra Trees Classifier (ET)); and advanced models (i.e. multi-layer perceptron (MLP), support vector machines (SVM)) ^10 17 18^. Ensemble models enhance prediction by learning patterns from large datasets, supporting medical decision-making and improving healthcare outcomes.

Early identification of individuals at high risk for CVD is crucial for timely treatment and saving lives ^11^. The ML models are pivotal to CVD risk prediction through capturing complex interactions among risk factors, outperforming traditional models like FRS, PCEs, and SCORE ^19^. Unlike conventional CVD risk prediction calculators, certain machine learning methods can better accommodate non-linear relationships, high-dimensional feature spaces, and, in some cases, missing data ^20^. They integrate real-world datasets, including wearable device data and electronic health records, continuously updating for improved accuracy ^21^. Improved ML-based prediction models are essential for Africa, ensuring better CVD risk assessment and tailored treatment, ultimately enhancing healthcare outcomes and saving lives. With the availability of CVD data through the WHO STEPWise Surveys (STEPS) in Africa, there is a need to predict CVD risk using ML methods in Africa between 2014 and 2019 ^22^. In this paper, we aim to apply the ML techniques on socio-behavioral characteristics by comparing the three different ML algorithms. This information will better equip the researchers and policymakers to analyze the CVD landscape in Africa, identify critical patterns and trends, and inform the development of targeted strategies, like uptake of CVD prophylaxis and treatment, for the individuals who are at most risk of CVDs in Africa or other similar settings.

## METHODS

### Data source

This analysis utilized the WHO STEPS with CVD data collected between 2014 and 2019 in Africa from the following twelve countries: Algeria, Benin, Botswana, Eswatini, Ethiopia, Kenya, Malawi, Morocco, São Tomé and Príncipe (STP), Sudan, Uganda, and Zambia. The WHO STEPS gather the individual-level demographic and behavioural risk factors, as well as the biomarkers, for non-communicable diseases using a multi-stage cluster sampling process. The behavioral risk factors like tobacco use, alcohol use, and unhealthy diet, as well as biological risk factors like overweight, obesity, blood pressure, glucose, and abnormal blood lipids ^23^.

We merged WHO STEPS data from 12 countries, standardizing categorical variables like residence, gender, occupation, and education for consistency. The dataset, containing 60,294 records, aimed to predict whether individuals had ever experienced a heart attack, chest pain, or stroke. Missing data were simulated using binomial, multinomial, and Gaussian distributions due to issues with the Multivariate Imputation by Chained Equations (MICE) method ^24^, which was unsuitable due to auto-correlation in data from twelve countries ^25^. This simulation approach ensured data completeness while preserving sample size for analysis ^26^.

### Variables definitions

Box 1 displays the variables that were part of the analysis. These were chosen because they were accessible in each of the 12 study countries. Self-reported CVD status/history, as indicated by a “yes” or “no” response to a question regarding a prior heart attack, angina, or stroke, was the outcome variable. In addition to age, sex, education, and marital status, predictor variables included biological, behavioural, and demographic characteristics such as blood pressure, physical activity, problematic alcohol use, and tobacco use. This thorough approach guarantees that important risk and preventive factors for CVD in a variety of populations are included in the analysis.

### CVD status imbalance treatment

To address the pronounced imbalance between CVD and non-CVD cases, we applied the Synthetic Minority Over-Sampling Technique (SMOTE), optimizing parameters (K=5, duplication size=2) to achieve an equitable class distribution. We compared SMOTE with alternative approaches, including ROSE, random under-sampling (RUS), and combined strategies, identifying SMOTE as the most effective. This adjustment markedly improved model sensitivity, raising it from below 8% to over 60% across all machine-learning algorithms, ensuring fairer and more reliable risk prediction.

### Machine-learning algorithms

The study partitioned the WHO STEPS data into a training set (80%) and an independent test set (20%) to evaluate the performance of machine learning algorithms ^27^ for CVD risk prediction. Cross-validation was applied within the training data to tune hyperparameters, compare algorithms and void overfitting during selection. Elastic Net logistic regression, random forest (RF), and XGBoost were selected for their effectiveness in prediction accuracy and interpretability^19 28^. The models were developed in RStudio using the caret ^29^, Boruta ^30^, glmnet ^31^, ranger ^32^, and xgboost ^33^ packages. Hyperparameters including regularization and penalty parameters for logistic regression, and tree depth, number, and learning rate for tree-based models were optimized using grid search.

To improve the performance of ML models, the “Boruta” R package was used to address feature multi-collinearity by selecting the most relevant features for predictive modeling ^30^. Boruta applies a random forest-based feature selection method, removing insignificant features iteratively. Predictive metrics, such as accuracy (correct predictions / total predictions), sensitivity (true positives / actual positives), specificity (true negatives / actual negatives), and F1 score (harmonic mean of the sensitivity and positive predictive value), were used to assess model performance ^10 34 35 36^. 5-fold cross-validation helped avoid overfitting and inflated metrics, ensuring robust model evaluation.

### Validation Strategy

Robust model validation was implemented using multiple complementary approaches. Stratified splitting preserved class proportions across training and testing sets, while geographic validation leveraged reserved countries to evaluate model generalizability across regions. Temporal validation, using year-based data splits, assessed stability over time. Additionally, 5-fold stratified cross-validation within the training set enabled robust hyperparameter tuning and minimized overfitting, collectively ensuring that model performance estimates were reliable and generalizable to diverse African contexts.

### Statistical analysis

All statistical analyses were performed using R (version 4.4.2, R Foundation for Statistical Computing, Vienna, Austria) within the RStudio environment (version 2024.4.2.764). Descriptive statistics were reported as counts and percentages for categorical variables, and medians with interquartile ranges for continuous variables. Machine learning models were trained on one dataset and validated on a separate set. The Boruta algorithm was used to identify key predictors of CVD, and standard errors with 95% confidence intervals were estimated using robust jackknife procedures.

Given the imbalanced nature of CVD outcomes, the area under the precision-recall curve (AUC-PR) served as the primary metric. Model performance was further evaluated using balanced accuracy, F1-score, sensitivity, and specificity to capture multiple dimensions of predictive ability. Statistical comparisons were conducted using DeLong tests for AUC differences, and calibration was assessed with Hosmer-Lemeshow tests and visual calibration plots, ensuring both discrimination and probability alignment were rigorously quantified. To improve classification performance, an optimal threshold for binary prediction was selected based on the point that maximized the F1-score, rather than using the conventional 0.5 cut-off^37^. This approach provided a better balance between precision and recall. Final binary outcomes were derived from predicted probabilities using this best threshold. Model performance was assessed using accuracy, precision, sensitivity, specificity, and the F1-score ^38 39 40^. Model performance was benchmarked against standard clinical approaches, including logistic regression baselines, as well as Framingham and WHO/ISH risk scores adapted for the African context. Machine-learning models demonstrated statistically significant improvements in discrimination, calibration, and overall predictive utility, highlighting the added value of data-driven, locally informed approaches over conventional clinical risk tools for cardiovascular risk stratification in African populations.

Lastly, predicted probability plots stratified by age group, sex, and hypertension status were generated to visualize patterns in CVD risk across subpopulations. Hypertension is a well-established and modifiable risk factor for CVD, making its inclusion in predictive models both clinically and epidemiologically sound ^41 42^. By incorporating hypertension status, the models can capture a key driver of CVD risk that interacts with demographic variables such as age and sex and this enhances the model’s ability to reflect real-world risk patterns ^43^.

## RESULTS

### Descriptive statistics

Amongst 60,294 individuals, the prevalence of CVD was 5%. The characteristics of the study population are shown in Table 1. The study population consisted of 60,294 individuals, with a majority (44.1%) aged 15-29 years and only 6.0% aged 60 years or older. Males (51.2%) were slightly more than females (48.8%), and most participants (59.7%) resided in rural areas. Education levels varied, with 29.0% having no formal education and only 10.2% attaining tertiary education. Self-employment was the most common occupation (47.3%), while 68.5% were currently married. Ethiopia (23.4%) and Algeria (21.1%) contributed the largest proportions of participants. The prevalence of low fruit (81.3%) and vegetable intake (53.7%), high salt intake (33.2%), and low physical activity levels (16.4%). Hypertension (8.5%), diabetes (2.6%), and high cholesterol (6.2%) were relatively low, while harmful alcohol use (2.9%) and smoking (10.2%) were also uncommon. Most participants had a normal BMI (53.0%) while 14.0% were classified as obese.

**Table 1:** Characteristics of the study population in the 12 countries in sub-Saharan Africa between 2014 and 2019.

| Characteristics | Study population |  |
| --- | --- | --- |
|  | n | weighted % |
| Total | 60294 | 100.0 |
| **Age group** |  |  |
| 15-29 | 19351 | 44.1 |
| 30-39 | 15853 | 23.0 |
| 40-49 | 11563 | 16.6 |
| 50-59 | 7929 | 10.3 |
| 60+ | 5598 | 6.0 |
| **Sex** |  |  |
| Female | 37010 | 48.8 |
| Male | 23284 | 51.2 |
| **Area of residence** |  |  |
| Rural | 33718 | 59.7 |
| Urban | 26576 | 40.3 |
| **Highest level of education** |  |  |
| None | 20174 | 29.0 |
| Primary | 19647 | 35.0 |
| Secondary | 14915 | 25.7 |
| Tertiary | 5558 | 10.2 |
| **Occupation** |  |  |
| Government employee | 4413 | 7.8 |
| Non-Govt employee | 4622 | 8.7 |
| Non-paid/Retired | 25400 | 36.2 |
| Self-employed | 25859 | 47.3 |
| **Marital status** |  |  |
| *Currently married* | 39949 | 68.5 |
| *Formerly married* | 4019 | 5.4 |
| *Never married* | 16326 | 26.1 |
| **Country** |  |  |
| *Algeria* | 6955 | 21.1 |
| *Benin* | 5115 | 1.8 |
| *Botswana* | 3888 | 0.6 |
| *Eswatini* | 3026 | 0.2 |
| *Ethiopia* | 9241 | 23.4 |
| *Kenya* | 4477 | 14.1 |
| *Malawi* | 4186 | 6.4 |
| *Morocco* | 4991 | 0.0 |
| *STP* | 2418 | 0.2 |
| *Sudan* | 7722 | 16.2 |
| *Uganda* | 3973 | 10.0 |
| *Zambia* | 4302 | 6.1 |
| **Having diabetes** |  |  |
| *No* | 58800 | 97.4 |
| *Yes* | 1494 | 2.6 |
| **Having high cholesterol level** |  |  |
| *Yes* | 5926 | 6.2 |
| *No* | 54368 | 93.8 |
| **Having hypertension** |  |  |
| *No* | 52772 | 91.5 |
| *Yes* | 7522 | 8.5 |
| **High salt intake** |  |  |
| *No* | 40961 | 66.8 |
| *Yes* | 19333 | 33.2 |
| **Low fruit uptake** |  |  |
| *No* | 14037 | 18.7 |
| *Yes* | 46257 | 81.3 |
| **Low vegetable uptake** |  |  |
| *No* | 31130 | 46.3 |
| *Yes* | 29164 | 53.7 |
| **Physical activity level** |  |  |
| *Low Level* | 12325 | 16.4 |
| *Moderate level* | 12725 | 19.7 |
| *High level* | 35244 | 63.9 |
| **Smoking history** |  |  |
| *Current* | 5047 | 10.2 |
| *Never* | 51899 | 83.2 |
| *Previous* | 3348 | 6.7 |
| **Harmful alcohol use** |  |  |
| No | 58491 | 97.1 |
| Yes | 1803 | 2.9 |
| **BMI** |  |  |
| <18.5 | 7899 | 15.5 |
| 18.5-24 | 29966 | 53.0 |
| 25-29 | 12417 | 17.5 |
| 30+ | 10012 | 14.0 |
| **Survey year** |  |  |
| 2014 | 10887 | 10.8 |
| 2015 | 18833 | 39.2 |
| 2016 | 14677 | 37.3 |
| 2017 | 13479 | 12.5 |
| 2019 | 2418 | 0.2 |

### Class Distribution Before and After SMOTE

Figure 1 highlights a pronounced shift in class distribution following the application of the Synthetic Minority Over-Sampling Technique. In the original dataset of 60,294 observations, individuals without cardiovascular disease dominated the sample, with 57,399 cases representing 95.2 percent, while only 2,895 individuals, or 4.8 percent, were classified as having CVD. After applying SMOTE, the total sample size nearly doubled to 114,798 observations. The number of CVD cases increased sharply from 2,895 to 57,399, representing a substantial artificial expansion of the minority class, while the number of non-CVD cases remained unchanged at 57,399. As a result, the post-SMOTE dataset achieved an exact 50:50 balance between CVD and non-CVD outcomes. This redistribution corrected the severe class imbalance observed in the original data and created a more equitable training set, enabling the models to better learn patterns associated with CVD and improving the robustness and fairness of risk prediction across African populations.

**Figure 1.**
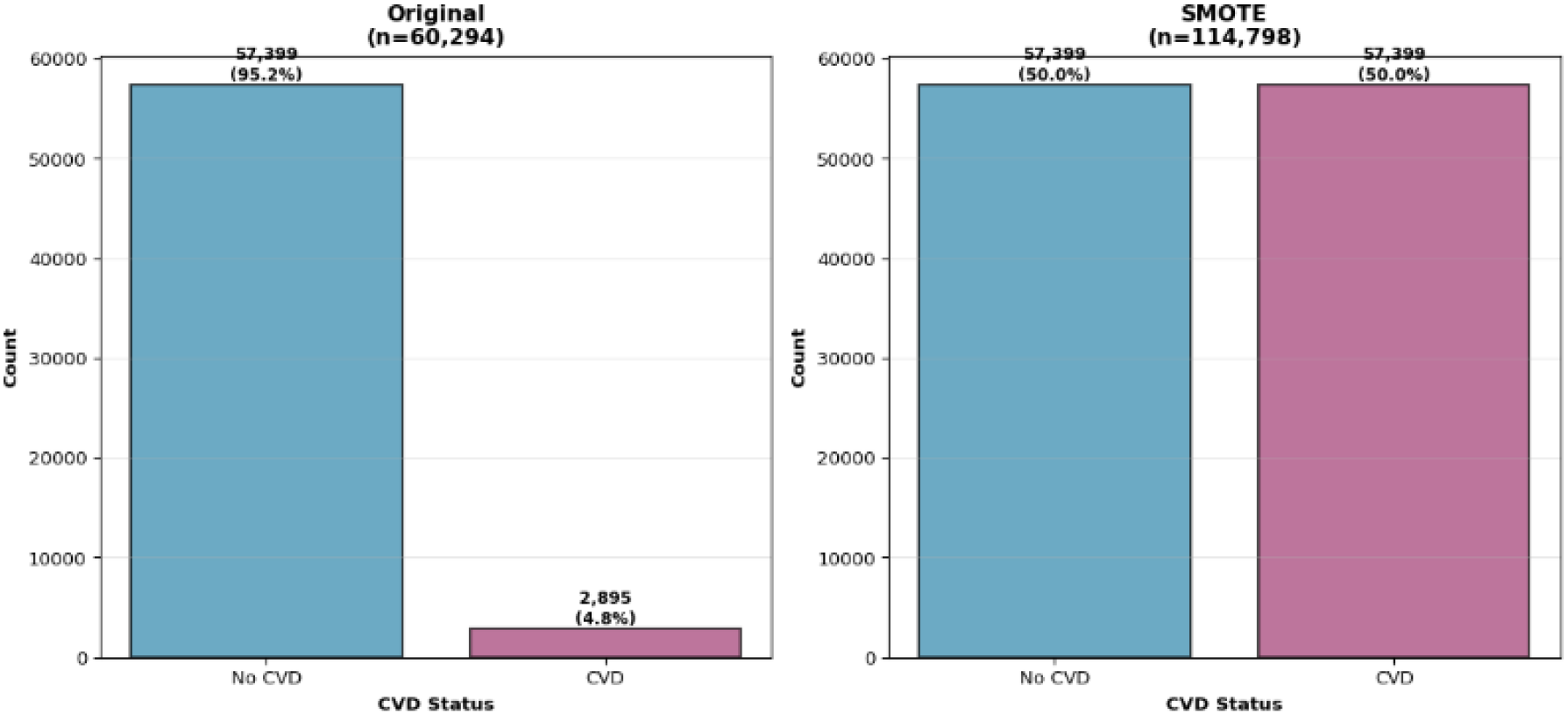
Data Distribution Before and After Synthetic Minority Over-Sampling Technique (SMOTE) for the Status of Cardiovascular Diseases in Africa (2014-2019)

### Feature importance and model explainability

Figure 2 shows a clear and intuitive pattern in how the models identified and ranked key predictors of cardiovascular disease risk across African STEPS data from 2014 to 2019. Blood pressure status emerged as the most influential feature by a wide margin, consistently dominating importance scores across all three models, which reinforces its central role in CVD risk in African populations. Alcohol-related harm was the second strongest predictor, highlighting the growing contribution of harmful alcohol use to cardiovascular risk in the region. Temporal and contextual factors such as survey year and country also ranked highly, suggesting meaningful variation in risk profiles across time and settings. Established clinical and behavioural factors including smoking history, diabetes, sex, and age category showed moderate importance, supporting their continued relevance in risk stratification. Dietary and lifestyle factors such as low fruit and vegetable intake, physical activity, and salt consumption contributed smaller but non-negligible effects, while socio-demographic variables including education, occupation, residence type, and marital status had relatively lower influence.

**Figure 2.**
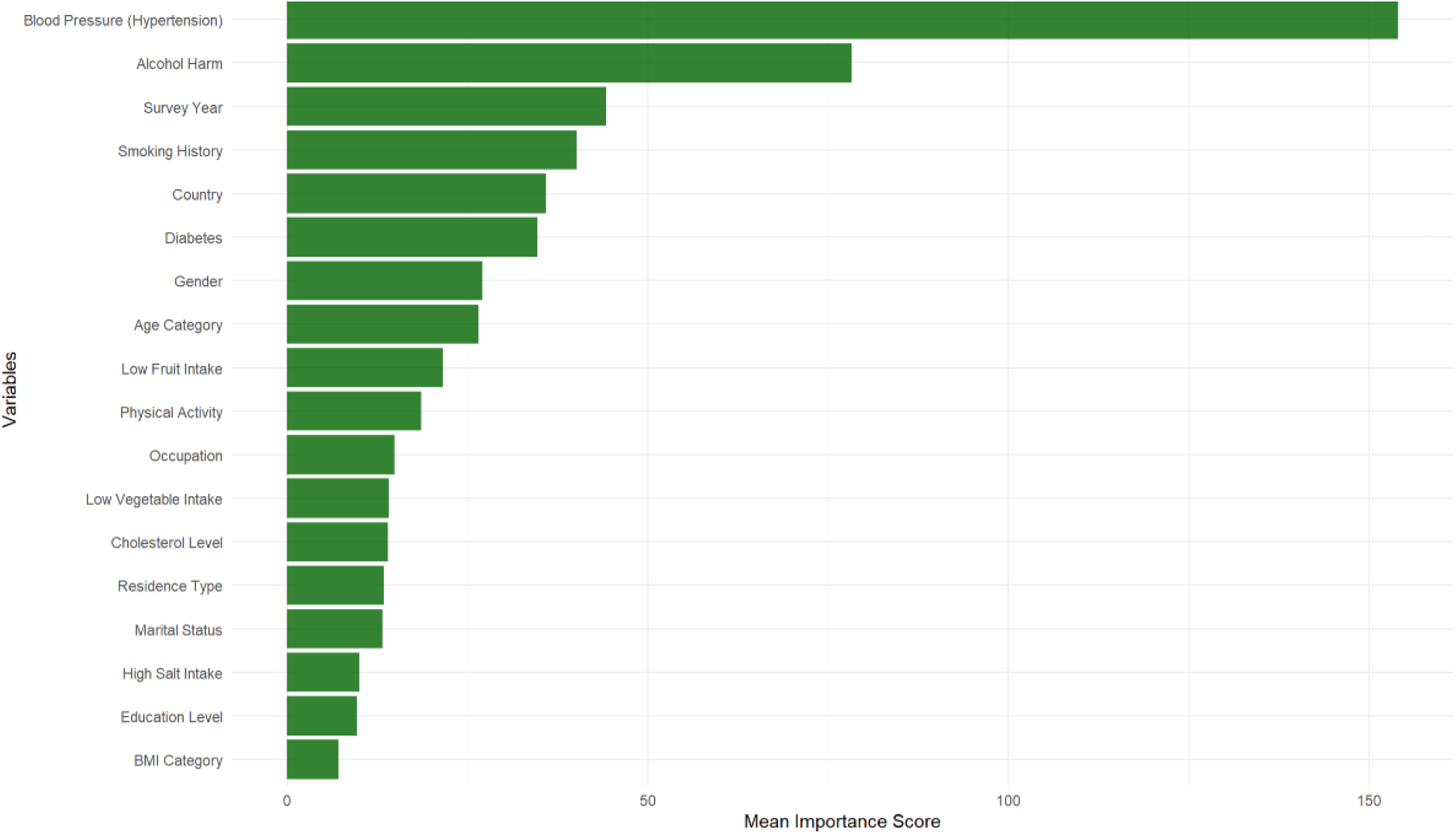
Features selected by the Boruta algorithm and their importance across three models after 5-fold cross-validation in Africa (2014-2019)

### Machine learning model performance

Table 2 highlights marked differences in predictive performance across the evaluated models. Among the machine learning approaches, XGBoost emerged as the strongest performer, achieving the highest discrimination with an AUC-ROC of 0.769, alongside a balanced accuracy of 0.699 and an F1 score of 0.710. It also showed a good balance between sensitivity at 0.729 and specificity at 0.668, with the lowest Brier score of 0.195, indicating better calibration and lower prediction error. Random Forest performed comparably, with an AUC-ROC of 0.758, balanced accuracy of 0.693, and F1 score of 0.699, although with slightly higher prediction error. In contrast, LASSO and logistic regression demonstrated weaker discrimination, with AUC-ROC values of 0.616 and 0.623, respectively. While LASSO achieved high sensitivity at 0.804, this was offset by very low specificity at 0.416, indicating substantial misclassification of non-CVD cases. Overall, these results suggest that tree-based machine learning models were better suited to capturing the complex risk patterns observed in the African STEPS data.

**Table 2:** Performance metrics for predictive modelling of prevalence of cardiovascular diseases in Africa (2014-2019)

| Model | AUC-ROC | AUC-PR | Balanced Accuracy | F1 Score | Sensitivity (Recall) | Specificity | Brier Score |
| --- | --- | --- | --- | --- | --- | --- | --- |
| **Machine Learning Models** |  |  |  |  |  |  |  |
| XGBoost | 0.769 | 0.350 | 0.699 | 0.710 | 0.729 | 0.668 | 0.195 |
| Random Forest | 0.758 | 0.353 | 0.693 | 0.699 | 0.707 | 0.678 | 0.204 |
| LASSO Regression | 0.616 | 0.453 | 0.610 | 0.676 | 0.804 | 0.416 | 0.239 |
| Logistic Regression | 0.623 | 0.437 | 0.604 | 0.650 | 0.725 | 0.484 | 0.238 |
| **Clinical Risk Scores** |  |  |  |  |  |  |  |
| WHO/ISH Risk Chart | 0.500 | 0.496 | 0.500 | 0.502 | 0.500 | 0.500 | 0.305 |
| Framingham Risk Score | 0.500 | 0.496 | 0.500 | 0.502 | 0.500 | 0.500 | 0.398 |

A pronounced performance gap was observed when comparing machine learning models with traditional clinical risk scores. Both the WHO/ISH risk chart and the Framingham risk score performed no better than chance, with identical AUC-ROC values of 0.500, balanced accuracy of 0.500, and equal sensitivity and specificity at 0.500. Their higher Brier scores, 0.305 for WHO/ISH and 0.398 for Framingham, further indicate poorer calibration and greater prediction error. In contrast, all machine learning models achieved substantially higher discrimination, with AUC-ROC values ranging from 0.616 to 0.769, and consistently better balance between sensitivity and specificity. These findings underscore the limitations of applying conventional clinical risk tools developed in high-income settings to African populations and highlight the added value of region-specific, data-driven machine learning models for cardiovascular disease risk prediction.

### Model calibration

Figure 3 shows clear and practically meaningful differences in how well cardiovascular risk prediction models align predicted risk with observed outcomes in African populations. Among all models assessed, XGBoost demonstrated the strongest calibration, with predicted probabilities tracking closely along the 45-degree reference line across most of the risk spectrum. This near-perfect alignment indicates minimal systematic error, with observed event rates increasing proportionally as predicted risk rose from approximately 10% to over 80%, even in larger samples approaching 12,000 participants. Such consistency suggests that XGBoost not only discriminates risk well but also translates predictions into clinically interpretable probabilities.

**Figure 3.**
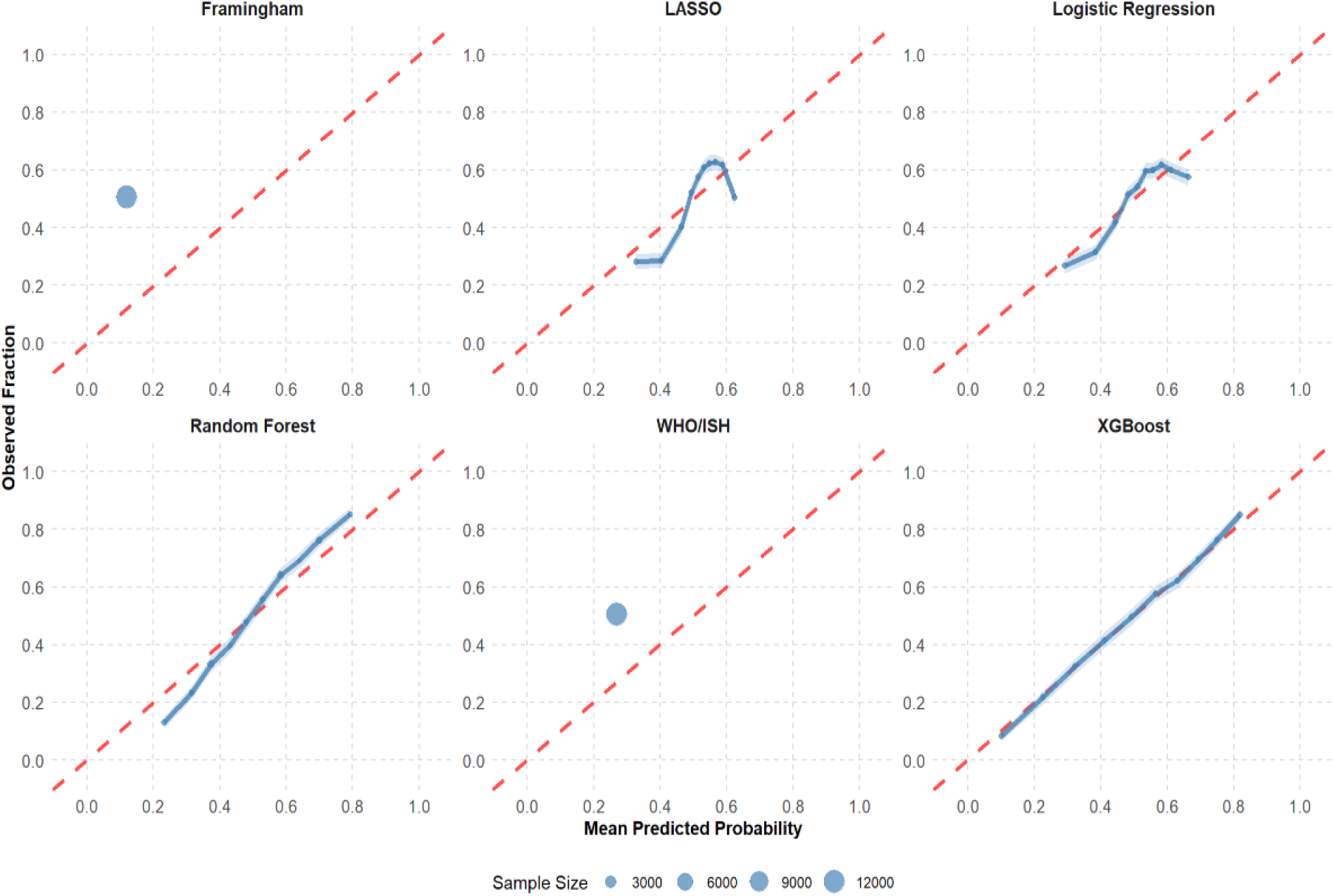
Model Calibration Assessment for Machine Learning and Clinical Decision Models of Cardiovascular Disease Risk in Africa (2014-2019)

In contrast, Random Forest models showed good but less uniform calibration, performing best in the moderate-to-high risk range (approximately 30–70% predicted risk). At lower predicted probabilities, Random Forest tended to underestimate observed risk, while slight overestimation emerged at higher risk levels. LASSO and conventional logistic regression models demonstrated acceptable calibration in the mid-range of predicted risk but deviated more noticeably at the extremes, where observed risks were either higher or lower than predicted. These departures suggest that simpler linear models may struggle to fully capture non-linear risk patterns present in African datasets.

Traditional clinical risk scores performed substantially worse. Framingham and WHO/ISH models showed marked miscalibration, with observed event rates clustering around 50% despite predicted risks below 20–30%, indicating systematic underprediction. This mismatch highlights the limited transportability of these scores to African populations and reinforces concerns that reliance on unadapted clinical risk charts may lead to under-identification of high-risk individuals in routine care.

### Predicted probability of reporting cardiovascular diseases

Across models, age and hypertension emerge as dominant and interrelated drivers of cardiovascular risk, but the magnitude, smoothness, and plausibility of predicted probabilities vary substantially by modelling approach. Machine-learning models; particularly XGBoost and LASSO; produce more conservative, gradual, and clinically intuitive risk trajectories, while traditional clinical scores show abrupt late-life risk inflation. These contrasts reinforce the importance of context-specific modelling and support the use of machine-learning approaches for more realistic cardiovascular risk estimation in African populations. The predicted probabilities of the specific models based on Figure 4 are presented below.

**Figure 4.**
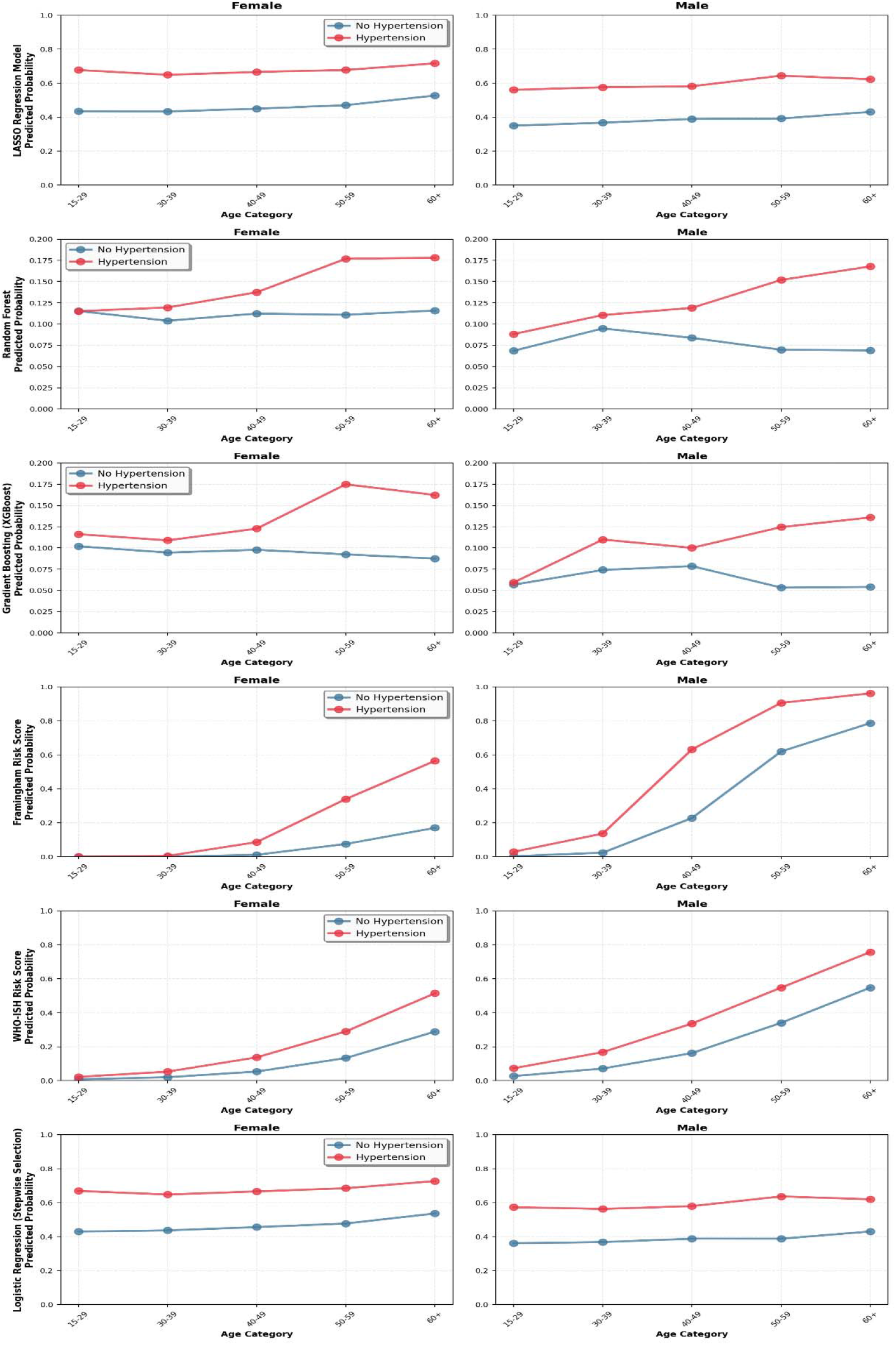
Predicted probabilities of developing cardiovascular diseases by age, sex and hypertension status from all the models in Africa (2014-2019)

#### LASSO regression

Figure 4 shows that the LASSO model consistently identifies hypertension as a strong and persistent amplifier of cardiovascular disease (CVD) risk across the life course, with age exerting a clear, monotonic effect. Overall, predicted probabilities increase steadily with advancing age, and at every age category, individuals with hypertension exhibit substantially higher predicted risk than their normotensive counterparts. Among males, the predicted probability for those with hypertension rises from approximately 12–13% in the 15–29-year age group to nearly 15–16% among those aged 60 years and above, while non-hypertensive males remain below 6% across all age categories, showing only modest age-related increases. This widening gap with age highlights the cumulative burden of hypertension on cardiovascular risk in men. A similar but slightly attenuated pattern is observed among females. Hypertensive women experience a steady increase in predicted risk from roughly 9–10% in early adulthood to over 12–13% in the oldest age group, whereas non-hypertensive women consistently exhibit lower probabilities, remaining under 6% throughout (see Figure 4). Collectively, these findings underscore the robust and stable influence of hypertension within a parsimonious linear framework, with minimal sex-based divergence but a clear age-dependent escalation of risk.

#### Random Forest

The Random Forest model presents a more nuanced picture, emphasizing age as the dominant driver of predicted CVD risk, while the modifying effect of hypertension appears less pronounced overall (see Figure 4). Across both sexes, predicted probabilities increase with age, particularly after midlife, suggesting the accumulation of multiple interacting risk factors in older populations. Among males, predicted probabilities rise sharply after age 50, peaking at approximately 18–19% in the 60+ age group for both hypertensive and non-hypertensive individuals. The convergence of risk estimates in older men implies that, within this model, hypertension may be overshadowed by other age-related factors, such as metabolic or lifestyle risks, once individuals reach older ages. In females, risk increases more gradually across age groups, with predicted probabilities ranging from around 11–12% in younger adults to approximately 17–18% in those aged 60 years and above. Interestingly, differences by hypertension status are modest, and in some age groups, normotensive women exhibit comparable or slightly higher predicted risk than hypertensive women (see Figure 4). This pattern suggests that Random Forest captures complex non-linear interactions, potentially diluting the isolated effect of hypertension, particularly among women.

#### XGBoost (Gradient Boosting)

The Gradient Boosting (XGBoost) model provides a balanced and clinically intuitive representation of cardiovascular risk, clearly integrating age, sex, and hypertension status (se Figure 4). Overall, predicted probabilities increase steadily with age in both sexes, and hypertension consistently confers an elevated risk across all age categories, reinforcing its role as a key independent risk factor. Among males, predicted CVD risk rises from approximately 6–7% in the youngest age group to nearly 18% among those aged 60 years and above. Hypertensive males consistently show higher probabilities than normotensive males at every age, although the absolute difference narrows slightly in older age groups as baseline risk increases. In females, a parallel trend is observed, with predicted probabilities increasing from around 7–8% in early adulthood to over 17–18% in the oldest age category among those with hypertension. Normotensive women maintain lower predicted risks throughout, though the gap again narrows with age (see Figure 4). Notably, sex differences are more evident at younger ages; where hypertensive men exhibit higher predicted risk; but largely converge or even slightly reverse in later life, reflecting differential risk accumulation across the life course.

#### Framingham risk score

The Framingham model produces markedly steeper age gradients than the machine-learning models, with predicted probabilities escalating rapidly in older age groups (see Figure 4). Overall, the model assigns very low risk in younger adults, followed by abrupt increases after age 40, particularly among individuals with hypertension. Among males, predicted probabilities for hypertensive individuals rise sharply from below 10% in younger age groups to nearly 90–95% by age 60 years and above, while non-hypertensive males also experience a dramatic increase, reaching approximately 75– 80% in the oldest age group. This pronounced separation suggests strong age- and hypertension-driven risk amplification, but may also indicate risk overestimation in older African populations. In females, the pattern is similar but less extreme. Hypertensive women experience a rise from near-zero predicted risk in early adulthood to approximately 55–60% in the oldest age group, while normotensive women reach only 15–20% (see Figure 4). The stark contrast between sexes and the abrupt risk escalation highlight the limited calibration and transferability of Framingham estimates in African settings.

#### WHO/ISH risk score

The WHO/ISH model shows progressive increases in predicted CVD risk with age, but with overall probabilities lower than those generated by Framingham (see Figure 4). Hypertension remains a key differentiator across age groups, although risk gradients are smoother. Among males, predicted probabilities increase from approximately 5–7% in young adults to 70–75% among hypertensive individuals aged 60 years and above, compared with around 50–55% for their non-hypertensive counterparts. This suggests substantial age-related escalation, with hypertension contributing meaningfully to risk stratification. In females, predicted risk rises from near 2–3% in early adulthood to over 50% among hypertensive women aged 60+, while normotensive women reach approximately 30%. Although less extreme than Framingham, the WHO/ISH model still assigns substantially higher late-life risks than machine-learning approaches, raising concerns about potential overestimation in older African women.

#### Logistic regression (stepwise selection)

The stepwise logistic regression model yields smooth, stable, and sex-consistent risk trajectories, closely resembling those observed in LASSO but with slightly higher baseline probabilities (see Figure 4). Overall, hypertension confers a clear upward shift in predicted risk at all ages, while age drives gradual increases rather than abrupt transitions. Among males, predicted probabilities for hypertensive individuals range from approximately 55–58% in younger age groups to over 60–63% in those aged 60+, compared with 35–40% among non-hypertensive males. The parallel trajectories suggest a persistent, additive effect of hypertension across the lifespan. Among females, hypertensive individuals show predicted probabilities increasing from roughly 60–62% to 65–68%, while normotensive women remain consistently lower, rising from about 45% to just over 50% (see Figure 4). The relatively narrow sex gap and smooth age gradients indicate stable model behaviour, though the higher absolute probabilities compared with tree-based models may reflect residual overprediction.

#### African Cardiovascular Disease Risk Prediction Tool

This African Cardiovascular Disease Risk Prediction Tool is an interactive, HTML-based decision-support application designed to estimate CVD risk in African populations using data from WHO STEPS surveys (2014–2019). It allows users to input basic patient characteristics; age group, sex, and blood pressure status; and compare predicted CVD risk across multiple approaches, including machine-learning models (LASSO, Random Forest, XGBoost, Logistic Regression) and established clinical risk scores (Framingham and WHO-ISH). The tool visually presents individual risk estimates, age-related risk progression, and cross-model comparisons, while applying clinically informed risk categorisation rules tailored to low-resource settings. Overall, it translates complex machine-learning outputs into an intuitive, clinically interpretable interface to support context-specific cardiovascular risk assessment, prevention planning, and policy discussions in Africa and similar settings.

## DISCUSSION

This study provides one of the most comprehensive evaluations to date of CVD risk prediction using machine-learning techniques in African populations, drawing on harmonised STEPS data from multiple countries between 2014 and 2019. Overall, we found that tree-based machine-learning models; particularly XGBoost and Random Forest; substantially outperformed both traditional regression approaches and widely used clinical risk scores in discriminating and calibrating CVD risk. Hypertension emerged as the single most influential predictor across all models, followed by alcohol-related harm, while age and sex shaped risk trajectories in expected but context-specific ways. In contrast, conventional clinical tools such as the Framingham and WHO/ISH risk scores showed poor discrimination and marked miscalibration, systematically underestimating risk at lower predicted probabilities and inflating risk in older age groups. Importantly, machine-learning models generated smoother, more plausible age- and sex-specific risk profiles, suggesting greater suitability for capturing the complex, non-linear risk patterns characteristic of African populations. Together, these findings highlight both the limitations of transplanting clinical risk scores developed in high-income settings and the potential of data-driven, region-specific machine-learning approaches to improve cardiovascular risk stratification and prevention efforts across Africa.

Globally, cardiovascular disease remains the leading cause of mortality, with risk accumulation often beginning decades before clinical disease becomes apparent. In high-income settings, declining CVD incidence has been driven largely by early detection and sustained risk-factor control^45^. In contrast, our finding of a relatively low self-reported CVD prevalence (5%) alongside extremely high levels of behavioural risk factors; such as poor diet, high salt intake, and physical inactivity; mirrors patterns reported across sub-Saharan Africa and other low-resource regions^46,47^. This apparent paradox reflects the younger age structure of African populations and substantial underdiagnosis, rather than a true absence of cardiovascular risk. From a policy perspective, these findings underscore the urgent need for primordial and primary prevention strategies, rather than reactive, treatment-focused approaches^48,49^. Programs that promote healthy diets, reduce salt intake, and encourage physical activity; embedded within schools, workplaces, and community settings; are likely to yield substantial long-term benefits. Importantly, the results highlight that waiting for clinically manifest CVD before intervening will miss a critical window for prevention in Africa.

Class imbalance is a common challenge in global health datasets, particularly for chronic diseases that remain underdiagnosed in low-income settings^50^. Our use of SMOTE to correct a severe imbalance between CVD and non-CVD cases aligns with best practices increasingly adopted in global machine-learning research^51^. Globally, failure to address such imbalance has been shown to bias models toward majority outcomes, disproportionately disadvantaging high-risk but underrepresented groups. In the African context, where CVD surveillance systems remain weak, correcting for imbalance is not merely a technical choice but an equity-driven methodological decision^52^. Programmatically, this highlights the importance of investing in better routine data systems and diagnostic capacity to reduce reliance on synthetic corrections. However, in the interim, balanced modelling approaches can help ensure that emerging digital risk tools do not systematically under-identify individuals at highest risk.

The central role of hypertension in driving CVD risk is well established globally, yet control rates remain particularly low in Africa, where awareness, treatment, and adherence are persistently suboptimal^53–55^. Our findings reinforce hypertension as the single most influential predictor of CVD across all models, consistent with evidence from both global pooled analyses and African cohort studies. Notably, the prominence of alcohol-related harm as the second strongest predictor reflects a growing but often underappreciated driver of cardiovascular risk in African settings. Policy implications are clear and actionable. Scaling up hypertension screening, affordable medication access, and long-term management within primary healthcare should remain a cornerstone of national NCD strategies^56^. At the same time, alcohol harm reduction; through taxation, marketing restrictions, Screening, Brief Intervention, and Referral to Treatment (SBIRT), and community-based interventions; must be elevated within cardiovascular prevention agendas^57,58^. These findings argue for integrated NCD policies that address both clinical and behavioural determinants rather than siloed interventions.

Globally, machine-learning models are increasingly shown to outperform conventional risk scores by capturing non-linear relationships and complex interactions among risk factors^59^. Our findings extend this evidence to African populations, demonstrating that tree-based models such as XGBoost and Random Forest substantially outperform both regression-based approaches and traditional clinical risk charts^60^. The poor performance of Framingham and WHO/ISH scores is consistent with prior African validation studies showing limited transportability of models developed in high-income contexts^61^. From a policy and programmatic standpoint, these results challenge the continued reliance on imported clinical risk tools in African guidelines. While ML models should not replace clinical judgment, they offer an opportunity to develop locally calibrated, digitally deployable risk stratification tools that can be integrated into routine care, mobile health platforms, and community screening programs^61^. Importantly, adoption should be accompanied by investments in transparency, clinician training, and ethical governance^62^.

Calibration is critical for translating predicted risk into real-world decision-making, yet it is often overlooked in global health modelling^63^. Our findings that XGBoost achieved the best calibration, while traditional scores showed marked miscalibration, are particularly important for African contexts where overtreatment and undertreatment both carry significant costs. Globally, poor calibration has been associated with inappropriate treatment thresholds and reduced clinician trust in risk tools. In African health systems with constrained resources, well-calibrated models are essential to prioritise limited preventive therapies, such as antihypertensive medications and statins, to those most likely to benefit. Programmatically, this supports the use of ML-based tools for risk stratification rather than universal treatment, improving efficiency while maintaining equity. However, routine recalibration using local data should be embedded into implementation strategies to maintain accuracy over time^64^.

Globally, sex differences in cardiovascular risk reflect a complex interplay of biology, behaviour, and social determinants. Our findings suggest that while overall sex differences in predicted risk were modest, important variations emerged by age and hypertension status, particularly in younger adults^65,66^. This aligns with African evidence showing earlier onset and poorer control of hypertension among men, alongside rising risk among older women as protective hormonal effects diminish. Policy implications include the need for sex-sensitive prevention strategies, such as targeted hypertension screening for younger men and improved cardiovascular risk assessment for post-menopausal women^67^. Programmes that assume uniform risk across sexes may fail to reach key subgroups, whereas ML-based tools offer the flexibility to capture and respond to these nuanced patterns.

The African Cardiovascular Disease Risk Prediction Tool represents a significant advancement in context-specific, data-driven cardiovascular risk assessment for African populations^68^. By leveraging harmonised WHO STEPS data across multiple countries and applying machine-learning techniques such as XGBoost and Random Forest, the tool overcomes key limitations of conventional risk scores developed in high-income settings, offering superior discrimination, calibration, and clinically interpretable risk estimates across age, sex, and hypertension status^69^. Its strength lies in capturing complex, non-linear interactions; particularly the variable influence of hypertension across demographic groups; producing smoother, more gradual risk trajectories than traditional tools and mitigating risks of over- or under-treatment. The incorporation of behavioural factors, including alcohol-related harm, further enhances its relevance for prevention-oriented strategies in African health systems^70^. Programmatically, the web-based design facilitates flexible deployment in clinical, community, and policy contexts, supporting targeted interventions without supplanting clinical judgment.

### Strengths and limitations

This study has several notable strengths. By leveraging harmonised WHO STEPS data from 12 African countries, it provides one of the most comprehensive, multi-country evaluations of CVD risk prediction on the continent. The application of advanced machine-learning models; particularly XGBoost and Random Forest; allowed the capture of complex, non-linear interactions between demographic, behavioural, and biological risk factors, producing more calibrated and clinically interpretable risk estimates than traditional tools such as Framingham and WHO/ISH scores^69^. Key predictors identified, including hypertension and alcohol-related harm, highlight actionable targets for prevention, supporting policy-relevant interventions such as scaling up hypertension detection and treatment programs, promoting lifestyle modification, and implementing alcohol harm reduction strategies like SBIRT^56^. The HTML-based African CVD Risk Prediction Tool further strengthens the study’s translational value, enabling flexible deployment in clinical, community, and policy settings to guide risk stratification and resource allocation.

However, several limitations warrant consideration. External validation is currently limited to African populations, leaving the generalizability of the models to other regions uncertain^71^. The models were trained on data collected between 2014 and 2019, and temporal shifts in epidemiology may affect future performance, emphasizing the need for ongoing recalibration^72^. Feature availability was constrained to variables collected in WHO STEPS surveys, excluding potentially informative clinical or biomarker data that could enhance predictive accuracy. Finally, the study did not assess the cost-effectiveness of implementing machine-learning–based risk prediction in routine care, an important consideration for resource-constrained health systems^73^. Despite these limitations, the findings provide a robust foundation for data-driven, context-specific cardiovascular risk assessment and targeted prevention strategies across Africa.

## CONCLUSION

In conclusion, this study demonstrates that machine-learning approaches, applied to harmonised African data, can provide more accurate, context-specific, and actionable estimates of cardiovascular risk than conventional clinical scores. By identifying key drivers such as hypertension and alcohol-related harm, the findings reinforce the importance of policy-aligned interventions, complementing frameworks like the WHO Package of Essential Noncommunicable Disease Interventions (PEN) and the HEARTS technical package, which emphasize early detection, risk stratification, and standardized hypertension management in primary care^74,75^. The African CVD Risk Prediction Tool translates complex data into an intuitive platform for clinicians, communities, and policymakers, supporting targeted prevention strategies. Looking ahead, next steps should include external validation across diverse African settings, integration of additional clinical and biomarker data, and evaluation of the cost-effectiveness and real-world impact of implementing such tools. Crucially, these efforts should remain people-centered, empowering healthcare workers, engaging communities, and fostering equitable access to prevention and treatment. By aligning innovative, machine-learning–driven approaches with established policy frameworks, Africa can take meaningful strides toward reducing the growing burden of cardiovascular disease and strengthening resilient, prevention-focused health systems.

## Supporting information

Supplementary item

## Data Availability

All data produced in the present work are contained in the manuscript

## ABBREVIATIONS

AB: AdaBoost
AUC: Area Under the Curve
AUC-PR: Area Under the Precision–Recall Curve
AUC-ROC: Area Under the Receiver Operating Characteristic Curve
BMI: Body Mass Index
Boruta: Boruta Feature Selection Algorithm
CVD: Cardiovascular Disease
DT: Decision Tree
ET: Extra Trees Classifier
FRS: Framingham Risk Score
GLMM: Generalized Linear Mixed Model
HICs: High-Income Countries
ISH: International Society of Hypertension
KNN: K-Nearest Neighbour
LASSO: Least Absolute Shrinkage and Selection Operator
LR: Logistic Regression
ML: Machine Learning
MLP: Multi-Layer Perceptron
NB: Naïve Bayes
NCD: Non-Communicable Disease
PCE: Pooled Cohort Equations
PEN: Package of Essential Noncommunicable Disease Interventions
QRISK3: Cardiovascular Disease Risk Algorithm Version 3
RF: Random Forest
ROSE: Random Over-Sampling Examples
RUS: Random Under-Sampling
SBIRT: Screening, Brief Intervention, and Referral to Treatment
SCORE: Systematic Coronary Risk Evaluation
SMOTE: Synthetic Minority Over-Sampling Technique
SSA: Sub-Saharan Africa
STEPS: STEPwise Approach to Surveillance
SVM: Support Vector Machine
WHO: World Health Organization
WHO/ISH: World Health Organization / International Society of Hypertension
XGB: Extreme Gradient Boosting
XGBoost: Extreme Gradient Boosting Machine

## Authors contributions

WFN performed data management, analysis and drafting the paper; AM, EO, JE and OK reviewed the analysis and the draft paper. All authors approved the final draft manuscript.

## Data availability

The datasets analysed during the current study were obtained from the WHO STEPwise Surveys and are not publicly available. Access to the raw data can be requested from the World Health Organization (WHO) STEPwise Surveillance Team through the WHO NCD Microdata Repository or by contacting the WHO NCD Surveillance Team at, subject to approval and relevant data-sharing agreements. The analytical code used in this study is available from the corresponding author upon reasonable request.

## Declarations

### Ethics approval and consent to participate

The WHO STEPS surveys were conducted in accordance with the ethical principles outlined in the Declaration of Helsinki. The WHO STEPwise Surveys included in this analysis received ethical approval from the relevant national institutional review boards or ethics committees in each participating country; Algeria, Benin, Botswana, Eswatini, Ethiopia, Kenya, Malawi, Morocco, São Tomé and Príncipe, Sudan, Uganda, and Zambia; as well as from the WHO Research Ethics Review Committee (ERC), World Health Organization. Written informed consent was obtained from all participants prior to data collection. The present study involved secondary analysis of anonymised data and did not require additional ethical approval.

This study is a secondary analysis of de-identified WHO STEPS survey data. The authors accessed the data through an approved data request to the World Health Organization. The analysis involved no direct contact with participants and did not require additional ethical approval, as the datasets were fully anonymized.

### Consent for publication

Not applicable. This manuscript does not contain any individual-level identifiable data.

---

**Box 1:** Outcome and predictor variables used in this analysis in Africa (2014-2019)

| Variable Type | Description | Categories |
| --- | --- | --- |
| **Outcome Variable** | "Have you ever had a heart attack or chest pain from heart disease (angina) or a stroke (cerebrovascular accident or incident)?" used as a proxy for CVD. | Yes/No |
| **Predictor Variables** | **Risk or protective factors for CVDs** |  |
| Current Tobacco Use | Smoking status | Current user, Previous user, Never used |
| Harmful Alcohol Use | Daily drinking with at least 4 (males) or 3 (females) drinking occasions in the last 30 days | Yes/No |
| Consumption of Vegetables | Frequency of vegetable intake of less than 5 days a week | Yes/No |
| Low Fruit Consumption | Frequency of fruit intake of less than 5 days a week | Yes/No |
| Physical Inactivity | Physical activity less than 150 minutes of moderate-intensity exercise per week | Yes/No |
| High Sugar Uptake | Regularly adding sugar to food, often eating processed foods with high sugar content, or no sugar intake | Yes/No |
| History of High Cholesterol | Previously diagnosed with high cholesterol | Yes/No |
| Road Traffic Accident | Involvement in a road traffic accident in the past 12 months | Yes/No |
| Violent Incident | Involvement in an incident where someone was injured and required medical attention | Yes/No |
| High Salt Intake | Regularly adding salt to food or often eating processed foods with high salt content | Yes/No |
| History of Raised Blood Pressure | Blood pressure $\geq 160/100$ mmHg or currently on medication | Yes/No |
| Systolic Blood Pressure | Systolic blood pressure (mmHg) | Continuous |
| Diastolic Blood Pressure | Diastolic blood pressure (mmHg) | Continuous |
| Fasting Blood Sugar | Fasting blood sugar levels (mmol/L) | Continuous |
| HDL Cholesterol | High-density lipoprotein cholesterol levels (mmol/L) | Continuous |
| History of Raised Blood Glucose | Capillary whole blood glucose $\geq 6.1$ mmol/L | Yes/No |
| Body Mass Index (BMI) | Categorized body mass index | <18.5, 18.5-24.0, 25.0-29.0, 30.0+ kg/m<sup>2</sup> (excluding pregnant women) |
| Age | Age of respondent (in completed years) | 15-29, 30-39, 40-49, 50-59, 60+ |
| Education | Highest level of education attained | None, Primary, Secondary, Tertiary |
| Occupation | Employment status | Government employee, Non-government employee, Self-employed, Retired/Not working (e.g., students) |
| Type of Residence | Place of residence | Rural, Urban |
| Sex | Sex of respondent | Male, Female |
| Country | Respondent country of residence | Malawi, Zambia, Tanzania, Kenya, etc. |
| Marital Status | Current marital status | Single, Married |

**Supplementary material for the paper: african_cvd_risk_prediction_tool.html**

An HTML-based tool to estimate cardiovascular disease risk using age, sex, and blood pressure, based on model-predicted probabilities.

## REFERENCES

1. Roth, G. A. et al. Global Burden of Cardiovascular Diseases and Risk Factors, 1990-2019: Update From the GBD 2019 Study. J. Am. Coll. Cardiol. 76, 2982–3021 (2020).

2. World Health Organization. WHO| Cardiovascular diseases (CVDs) [Internet]. Geneva: World Health Organization; 2017. http://www.who.int/cardiovascular_diseases/en/.

3. Kim, K.-I. Risk Stratification of Cardiovascular Disease according to Age Groups in New Prevention Guidelines: A Review. J. Lipid Atheroscler. 12, 96–105 (2023).

4. Global Burden of Disease Collaborative Network, Global Burden of Disease Study 2019 (GBD 2019) Results (2020, Institute for Health Metrics and Evaluation – IHME) https://vizhub.healthdata.org/gbd-results/.

5. Sazlina, S. G. et al. Cardiovascular disease risk factors among older people: Data from the National Health and Morbidity Survey 2015. PloS One 15, e0240826 (2020).

6. Minja, N. W. et al. Cardiovascular diseases in Africa in the twenty-first century: Gaps and priorities going forward. Front. Cardiovasc. Med. 9, 1008335 (2022).

7. Mroz, T. et al. Predicting hypertension control using machine learning. PloS One 19, e0299932 (2024).

8. Ahmad, A. A. & Polat, H. Prediction of Heart Disease Based on Machine Learning Using Jellyfish Optimization Algorithm. Diagn. Basel Switz. 13, (2023).

9. Chou, C.-Y.Hsu, D.-Y. & Chou, C.-H. Predicting the Onset of Diabetes with Machine Learning Methods. J. Pers. Med. 13, (2023).

10. Asadi, F. et al. Detection of cardiovascular disease cases using advanced tree-based machine learning algorithms. Sci. Rep. 14, 22230 (2024).

11. Johnson, A. R., Arasu, S. & Gnanaselvam, N. A. Cardiovascular Disease Risk Factors and 10 Year Risk of Cardiovascular Events among Women over the Age of 40 Years in an Urban Underprivileged Area of Bangalore City. J. -Life Health 12, 225–231 (2021).

12. Lloyd-Jones, D. M., Larson, M. G., Beiser, A. & Levy, D. Lifetime risk of developing coronary heart disease. The Lancet 353, 89–92 (1999).

13. Berger, J. S., Jordan, C. O., Lloyd-Jones, D. & Blumenthal, R. S. Screening for cardiovascular risk in asymptomatic patients. J. Am. Coll. Cardiol. 55, 1169–1177 (2010).

14. Stevens, D., Lane, D. A., Harrison, S. L., Lip, G. Y. H. & Kolamunnage-Dona, R. Modelling of longitudinal data to predict cardiovascular disease risk: a methodological review. BMC Med. Res. Methodol. 21, 283 (2021).

15. Hippisley-Cox, J., Coupland, C. & Brindle, P. Development and validation of QRISK3 risk prediction algorithms to estimate future risk of cardiovascular disease: prospective cohort study. bmj 357, (2017).

16. Nomali, M. et al. Validity of the models predicting 10-year risk of cardiovascular diseases in Asia: A systematic review and prediction model meta-analysis. PLOS ONE 18, e0292396 (2023).

17. Hajjem, A., Larocque, D. & Bellavance, F. Generalized mixed effects regression trees. Stat. Probab. Lett. 126, 114–118 (2017).

18. Polley, Eric C. and van Der Laan, Mark J., ‘Super Learner In Prediction’ (May 2010). U.C. Berkeley Division of Biostatistics Working Paper Series. Working Paper 266. Https://Biostats.Bepress.Com/Ucbbiostat/Paper266.

19. Weng, S. F., Reps, J., Kai, J., Garibaldi, J. M. & Qureshi, N. Can machine-learning improve cardiovascular risk prediction using routine clinical data? PloS One 12, e0174944 (2017).

20. Stahl, D. New horizons in prediction modelling using machine learning in older people’s healthcare research. Age Ageing 53, (2024).

21. Hippisley-Cox, J. et al. Development and validation of a new algorithm for improved cardiovascular risk prediction. Nat. Med. 30, 1440–1447 (2024).

22. Cho, S.-Y. et al. Pre-existing and machine learning-based models for cardiovascular risk prediction. Sci. Rep. 11, 8886 (2021).

23. WHO. STEPwise approach to NCD risk factor surveillance (STEPS). (2022).

24. Azur, M. J., Stuart, E. A., Frangakis, C. & Leaf, P. J. Multiple imputation by chained equations: what is it and how does it work? Int. J. Methods Psychiatr. Res. 20, 40–49 (2011).

25. Lee, K. & Simpson, J. Introduction to multiple imputation for dealing with missing data. Respirol. Carlton Vic 19, (2013).

26. Morris, T. P., White, I. R. & Crowther, M. J. Using simulation studies to evaluate statistical methods. Stat. Med. 38, 2074–2102 (2019).

27. Gholamy, Afshin; Kreinovich, Vladik; and Kosheleva, Olga, ‘Why 70/30 or 80/20 Relation Between Training and Testing Sets: A Pedagogical Explanation’ (2018). Departmental Technical Reports (CS). 1209. Https://Scholarworks.Utep.Edu/Cs_techrep/1209.

28. Zhang, M., Wang, H. & Zhao, J. Use machine learning models to identify and assess risk factors for coronary artery disease. PLOS ONE 19, e0307952 (2024).

29. Kuhn, M. Building Predictive Models in R Using the caret Package. J. Stat. Softw. 28, 1–26 (2008).

30. Kursa, M. B. & Rudnicki, W. R. Feature Selection with the Boruta Package. J. Stat. Softw. 36, 1–13 (2010).

31. Friedman, J. H., Hastie, T. & Tibshirani, R. Regularization Paths for Generalized Linear Models via Coordinate Descent. J. Stat. Softw. 33, 1–22 (2010).

32. Wright, M. N. & Ziegler, A. ranger: A Fast Implementation of Random Forests for High Dimensional Data in C++ and R. J. Stat. Softw. 77, 1–17 (2017).

33. Chen, T.Q. and Guestrin, C. (2016) Xgboost: A Scalable Tree Boosting System. Proceedings of the 22nd ACM SIGKDD International Conference on Knowledge Discovery and Data Mining, San Francisco, 13-17 August 2016, 785–794. 10.1145/2939672.2939785.

34. Yang, L. et al. Study of cardiovascular disease prediction model based on random forest in eastern China. Sci. Rep. 10, 5245 (2020).

35. Gareth James, D. W., Trevor Hastie, Robert Tibshirani. An Introduction to Statistical LearningfZ: With Applications in R. (New York[]: Springer, [2013] ©2013, 2013).

36. Cawley, G. C. & Talbot, N. L. C. On Over-fitting in Model Selection and Subsequent Selection Bias in Performance Evaluation. J Mach Learn Res 11, 2079–2107 (2010).

37. Lipton, Z. C., Elkan, C. & Naryanaswamy, B. Optimal Thresholding of Classifiers to Maximize F1 Measure. Mach. Learn. Knowl. Discov. Databases Eur. Conf. ECML PKDD Proc. ECML PKDD Conf. 8725, 225–239 (2014).

38. Cabot, J. H. & Ross, E. G. Evaluating prediction model performance. Surgery 174, 723–726 (2023).

39. Raschka, S. An Overview of General Performance Metrics of Binary Classifier Systems. https://doi.org/10.13140/2.1.4639.7440 (2014) doi:10.13140/2.1.4639.7440.

40. Powers, D. & Ailab. Evaluation: From precision, recall and F-measure to ROC, informedness, markedness & correlation. J Mach Learn Technol 2, 2229–3981 (2011).

41. Mills, K. T., Stefanescu, A. & He, J. The global epidemiology of hypertension. Nat. Rev. Nephrol. 16, 223–237 (2020).

42. Magnussen, C. et al. Global Effect of Modifiable Risk Factors on Cardiovascular Disease and Mortality. N. Engl. J. Med. 389, 1273–1285 (2023).

43. Oparil, S. et al. Hypertension. Nat. Rev. Dis. Primer 4, 18014 (2018).

44. Osei, E. et al. Prevalence and predictors of selected risk factors of non-communicable diseases in Ghana: evidence from a sub-national survey. J. Glob. Health Sci. 3, 1–14 (2021).

45. Lopez, A. & Adair, T. Is the long-term decline in cardiovascular-disease mortality in high-income countries over? Evidence from national vital statistics. Int. J. Epidemiol. 48, (2019).

46. Odunaiya, N., Grimmer, K. & Louw, Q. High prevalence and clustering of modifiable CVD risk factors among rural adolescents in southwest Nigeria: Implication for grass root prevention. BMC Public Health 15, 661 (2015).

47. Ng’ambi, W. F. et al. The Prevalence, Prevention, and Treatment of Cardiovascular Diseases in Twelve African Countries (2014-2019): An Analysis of the World Health Organisation STEPwise Approach to Chronic Disease Risk Factor Surveillance. medRxiv 2025.02.19.25322533 (2025) doi:10.1101/2025.02.19.25322533.

48. Phiri, N. et al. Development and piloting of a primary school-based salt reduction programme: Formative work and a process evaluation in rural and urban Malawi. PLOS Glob. Public Health 3, e0000867 (2023).

49. Silva-Santos, T. et al. Interventions That Successfully Reduced Adults Salt Intake—A Systematic Review. Nutrients 14, 6 (2022).

50. Salmi, M., Atif, D., Oliva, D., Abraham, A. & Ventura, S. Handling imbalanced medical datasets: review of a decade of research. Artif. Intell. Rev. 57, 273 (2024).

51. Sowjanya, A. M. & Mrudula, O. Effective treatment of imbalanced datasets in health care using modified SMOTE coupled with stacked deep learning algorithms. Appl. Nanosci. 13, 1829–1840 (2023).

52. Cau, R., Pisu, F., Suri, J. & Saba, L. Addressing hidden risks: Systematic review of artificial intelligence biases across racial and ethnic groups in cardiovascular diseases. Eur. J. Radiol. 183, 111867 (2024).

53. Govender, K. C. & Naidoo, M. Prevalence and determinants of apparent treatment-resistant hypertension among patients in South African primary care: a single-centre observational study. BMC Cardiovasc. Disord. 25, 373 (2025).

54. Gafane-Matemane, L. F. et al. Hypertension in sub-Saharan Africa: the current profile, recent advances, gaps, and priorities. J. Hum. Hypertens. 39, 95–110 (2025).

55. Brettler, J. W. et al. Drivers and scorecards to improve hypertension control in primary care practice: Recommendations from the HEARTS in the Americas Innovation Group. Lancet Reg. Health – Am. 9, (2022).

56. Dzudie, A. et al. Roadmap to Achieve 25% Hypertension Control in Africa by 2025. Glob. Heart 65, 105–110 (2019).

57. Gomez, E. et al. Using SBIRT (Screen, Brief Intervention, and Referral Treatment) Training to Reduce the Stigmatization of Substance Use Disorders Among Students and Practitioners. Subst. Abuse Res. Treat. 17, 11782218221146391 (2023).

58. Harris, L. M. et al. Teaching Note—SBIRT: Opportunities to Expand Addiction-Related Clinical Processes, Technical Skills, and Relational Competencies in Social Work. J. Soc. Work Educ. 1–16 doi:10.1080/10437797.2025.2466638.

59. Rahman, Y. & Dua, P. A machine learning framework for predicting healthcare utilization and risk factors. Healthc. Anal. 8, 100411 (2025).

60. Banerjee, M., Reynolds, E., Andersson, H. B. & Nallamothu, B. K. Tree-Based Analysis. Circ. Cardiovasc. Qual. Outcomes 12, e004879 (2019).

61. D’Agostino, R. B. S., Grundy, S., Sullivan, L. M. & Wilson, P. Validation of the Framingham coronary heart disease prediction scores: results of a multiple ethnic groups investigation. JAMA 286, 180–187 (2001).

62. Osonuga, A. et al. Bridging the digital divide: artificial intelligence as a catalyst for health equity in primary care settings. Int. J. Med. Inf. 204, 106051 (2025).

63. Alharbi, H. S. Interpretable and Calibrated XGBoost Framework for Risk-Informed Probabilistic Prediction of Slope Stability. Sustainability 17, (2025).

64. Ali, J. & Luqman, S. Using Machine Learning for Risk-based Monitoring and Data Integrity in Clinical Trials. (2023).

65. Sud, M. et al. Sex Differences in Cardiovascular Health Status and Long-Term Outcomes in a Primary Prevention Cohort. JACC Adv. 4, 102108 (2025).

66. Velásquez, I. et al. Sex Differences in the Relationship of Socioeconomic Position With Cardiovascular Disease, Cardiovascular Risk Factors, and Estimated Cardiovascular Disease Risk: Results of the German National Cohort. J. Am. Heart Assoc. 14, e038708 (2025).

67. Maas, A. et al. Changing clinical perspectives on sex and healthcare disparities in ischaemic heart disease. Lancet Reg. Health – Eur. 56, (2025).

68. Boateng, D. et al. Cardiovascular disease risk prediction in sub-Saharan African populations-Comparative analysis of risk algorithms in the RODAM study. Int. J. Cardiol. 254, (2018).

69. Kahraman, A. Machine learning techniques for improved prediction of cardiovascular diseases using integrated healthcare data. Front. Artif. Intell. 8, 1694450 (2025).

70. Ferreira-Borges, C., Dias, S., Babor, T., Esser, M. B. & Parry, C. D. H. Alcohol and public health in Africa: can we prevent alcohol-related harm from increasing? Addict. Abingdon Engl. 110, 1373–1379 (2015).

71. Arun, S. et al. Systematic scoping review of external validation studies of AI pathology models for lung cancer diagnosis. NPJ Precis. Oncol. 9, 166 (2025).

72. Patharkar, A., Cai, F., Al-Hindawi, F. & Wu, T. Predictive modeling of biomedical temporal data in healthcare applications: review and future directions. Front. Physiol. 15, 1386760 (2024).

73. El Arab, R. A. & Al Moosa, O. A. Systematic review of cost effectiveness and budget impact of artificial intelligence in healthcare. NPJ Digit. Med. 8, 548 (2025).

74. Brettler, J. W. et al. Drivers and scorecards to improve hypertension control in primary care practice: Recommendations from the HEARTS in the Americas Innovation Group. Lancet Reg. Health Am. 9, None (2022).

75. World Health Organization. WHO Package of Essential Noncommunicable (PEN) Disease Interventions for Primary Health Care. (World Health Organization, 2021).

